# Overview and evaluation of a nationwide hospital-based surveillance system for Influenza and COVID-19 in Switzerland (CH-SUR): 2018-2023

**DOI:** 10.1101/2024.09.18.24313869

**Authors:** Jonathan Aryeh Sobel, Marie-Céline Zanella, Rebecca Grant, Camille Beatrice Valera, Mária Suveges, Laura Urbini, Khaled Mostaguir, Sara Botero, Ursina Roder, Davide Bosetti, Rami Sommerstein, Ulrich Heininger, Petra Zimmermann, Peter W Schreiber, Domenica Flury, Anita Niederer-Loher, Philipp Jent, Alexia Cusini, Didier Pittet, Stephan Harbarth, Anne Iten, Olivia Keiser, the CH-SUR Collaborative Network

## Abstract

**Background:** In 2018, a hospital-based surveillance system for influenza (CH-SUR) was established in six tertiary care hospitals in Switzerland. From March 2020 onwards, this surveillance system was expanded to include more institutions, as well as COVID-19.

**Aim:** To quantitatively evaluate the timeliness and completeness of CH-SUR data and to qualitatively assess stakeholder perceptions of the importance, reliability and adaptability of the surveillance system.

**Methods:** All patients admitted to one of the participating centres for more than 24 hours and who had a laboratory-confirmed influenza virus or SARS-CoV-2 infection were included in CH-SUR. For all cases, we evaluated the timeliness and completeness of reporting to CH-SUR. A qualitative survey among CH-SUR stakeholders assessed perceived importance, understanding, reliability and adaptability of CH-SUR.

**Results:** Up to 20 centres participated in CH-SUR. Between December 2018 and October 2023, 7,375 cases of influenza were reported and between March 2020 and October 2023, 49,235 cases of COVID-19 were reported to CH-SUR. During the COVID-19 pandemic, time to data entry and completeness improved over time; the median delay of data entry in CH-SUR was 5 days (IQR=2-23) for COVID-19 and 4 days (IQR=2-15) for influenza during the period 2018-2023. The completeness of variables was high (99.4%), with the exception of COVID-19 or annual influenza vaccination status (respectively 15% and 72% of “Unknown” responses). Stakeholders perceived the system as important, relevant, understandable and adaptable.

**Conclusion:** CH-SUR has provided critical epidemiological and clinical information on hospitalised influenza and COVID-19 cases across Switzerland during the pandemic. Our evaluation highlighted the importance and relevance of this system among CH-SUR stakeholders, as well as its importance for preparedness and response to future infectious disease outbreaks.

## Introduction

Epidemiological surveillance systems are essential tools for monitoring and controlling the spread of diseases ^1,2,3,4^. In 2018, a hospital-based surveillance system for influenza (CH-SUR), jointly coordinated by Geneva University Hospitals (HUG), the Institute of Global Health at the University of Geneva (ISG UNIGE), and the Federal Office of Public Health (FOPH) was established in six tertiary care hospitals in Switzerland. The CH-SUR system was designed with three core objectives: to monitor in real time the clinical burden of hospitalised influenza and COVID-19 cases, to support efficient healthcare resource planning, and to evaluate the impact of public health interventions.

A standardised, electronic case report form (CRF) was established across all centres, using REDCap^5^. This enabled the continuous collection of demographics, clinical and epidemiological data from all patients admitted to one of the participating centres for more than 24 hours and who had a laboratory-confirmed influenza virus infection.

The first confirmed case of COVID-19 in Switzerland was reported to the FOPH on 25 February 2020 ^6^. As early as 2 March 2020, the CH-SUR system was expanded to include all patients admitted to one of the participating centres for more than 24 hours and who had a laboratory-confirmed SARS-CoV-2 infection. From 15 November 2020 onwards, the influenza and COVID-19 databases in CH-SUR were merged into a unified registry. The number of centres actively participating in CH-SUR increased during 2020, up to 20 hospitals and clinics, covering approximately 97% of all hospitalisations related to SARS-CoV-2 in Switzerland; 17 centres continued data collection until 2023 ^1–4,7,8^. (map of participating centres provided in Supplementary Figure S1).

Together with the primary care surveillance system (Sentinella) and community wastewater monitoring, CH-SUR served as one of the key pillars contributing to comprehensive surveillance of influenza and SARS-CoV-2 infections across Switzerland. The CH-SUR surveillance system was intended to enable Swiss health authorities to monitor epidemiological trends in hospitalised COVID-19 and influenza patients across Switzerland, to quantify the disease burden among hospitalised patients and to assess healthcare demands, thereby guiding the broader public health response to influenza and COVID-19, respectively. During the COVID-19 pandemic, key findings from the CH-SUR system were compiled into a monthly report published by HUG, ISG, and FOPH^9^ until September 2023.

To ensure that CH-SUR was meeting its stated objectives, an evaluation of CH-SUR was initiated by HUG, ISG, and FOPH. This study aimed to (i) quantify the timeliness and completeness of data entry in CH-SUR for influenza and COVID-19 cases, (ii) evaluate stakeholder perceptions of the system’s importance, reliability and adaptability, and (iii) identify key strengths and opportunities for improvement to guide future hospital-based surveillance.

## Methods

### Study design

We conducted a retrospective quantitative and qualitative evaluation of the nationwide hospital-based surveillance system for influenza and COVID-19 in Switzerland from 1 December 2018 to 17 October 2023 following CDC’s guidelines for evaluating public health surveillance systems^10^.

### Case definitions

All patients admitted to one of the participating centres for more than 24 hours and who had a laboratory-confirmed influenza virus infection or laboratory-confirmed or clinically diagnosed SARS-CoV-2 infection were included in CH-SUR. Patients who were seen in outpatient settings, or who were hospitalised for less than 24 hours were not included in CH-SUR. Within CH-SUR, cases were categorised as ‘nosocomial’ if a patient tested positive for influenza virus three days or more after hospital admission, or for SARS-CoV-2 five days or more after hospital admission. Cases transferred from other institutions were also considered as nosocomial cases.

Influenza cases were categorized by seasons, while COVID-19 cases were categorised by nine distinct phases, determined by the epidemiology and/or variant circulation during the pandemic: 1) Spring wave 2020 (24 February 2020 to 7 June 2020), 2) Fall/winter wave 2020 (8 June 2020 to 14 February 2021), 3) Alpha wave (15 February 2021 to 20 June 2021), 4) Delta wave (21 June 2021 to 19 December 2021), 5) BA.1 (Omicron) wave (20 December 2021 to 28 February 2022), 6) BA.2 (Omicron) wave (1 March 2022 to 5 June 2022), 7) BA.5 (Omicron) wave (6 June 2022 to 14 November 2022), 8) BQ.1 (Omicron) wave (15 November 2022 to 11 February 2023), and 9) XBB wave (12 February 2023 to 17 December 2023). The classification of SARS-CoV-2 variants was based on genomic sequencing data from Switzerland submitted to the Global Initiative on Sharing All Influenza Data (GISAID).^11^.

### Data entry and validation

The CH-SUR surveillance system involved multiple stakeholders (Supplementary Figure S2), including FOPH, ISG, the clinical research centre (CRC) of HUG and up to 20 hospitals across Switzerland. CH-SUR operated under the aegis of FOPH, which functioned as sponsor and project manager. Each of the participating centres had a principal investigator (PI). The PIs were responsible for the implementation of the surveillance system in their institution, as well as the collection and the quality of data; data collection was mostly performed by research nurses in each institution. ISG centralised data from CRFs for processing, evaluation and reporting. The ISG and the HUG CRC ensured the maintenance of the database, with HUG CRC also ensuring the hosting and security of the collected data.

All participating centres were required to complete an electronic CRF for all patients meeting the case definitions above. The CH-SUR CRF had six components: inclusion criteria, demographic information, case-related information, hospital admission data and clinical data throughout the course of the patient’s hospitalisation, including treatments received, admission to intensive or intermediate care units (ICU/IMCU) and outcome information. The CH-SUR codebook (version 2023) used during the data extraction is provided in the supplementary material (Supplementary File 1). The target for the time between patient admission to hospital (or positive test for nosocomial cases) and CH-SUR data entry for admission related information was 48 to 72 hours and the target for time to completion of data entry was 14 days. These targets were set by FOPH at the establishment of CH-SUR in 2018. The electronic CRF included internal consistency checks and pop-up messages for incomplete data entry. There was also a distinction between "Unknown" (information not available) and "NA" (missing data).

### Data extraction

For all influenza and COVID-19 cases in the CH-SUR database during the study period, we extracted the following variables: hospital admission date, date of positive influenza/SARS-CoV-2 test; COVID-19 or influenza vaccination information; ICU or IMCU admission data; outcome (in-hospital death, discharge alive). For COVID-19, the reason for hospitalisation (because of COVID-19 or with COVID-19) was introduced into the questionnaire in December 2021 “Because of COVID-19” refered to patients hospitalized primarily due to symptoms caused by COVID-19 or chronic conditions exacerbated by the virus, such as COPD or cardiac decompensation during infection. “With COVID-19” denoted patients hospitalized for reasons deemed to be unrelated to COVID-19 by the investigators but were SARS-CoV-2 test positive during their stay, for example hospitalisation for surgical procedures or infections other than SARS-CoV-2; this variable was also included in the data extraction. We also extracted data on the completeness of each of the six components of the CRF (Supplementary File 1). In the CRF, “NA” was used to denote fields that were not applicable for a given patient, while “Unknown” indicated that the information was unavailable at the time of data entry. For our quantitative analyses, we treated both “NA” and “Unknown” as missing values. Completeness for each variable was then calculated as the proportion of entries that were neither “NA” nor “Unknown.” We did not perform any imputation; all analyses of timeliness and completeness were conducted on available-case data only.

### Qualitative assessment

We conducted a REDCap survey among CH-SUR stakeholders (CH-SUR PIs, research nurses, data scientists/researchers, FOPH/ISG coordinators). These four groups supported the following CH-SUR activities: implementation of the surveillance system in participating centres, data collection, data quality assessments, clinical interpretation, reporting and coordination of the surveillance system. The survey questionnaire was designed to assess multiple dimensions of the CH-SUR project, including perceived importance, relevance, understanding and adaptability. A total of 67 questions were structured to elicit responses regarding the stakeholders’ perception of these aspects in relation to the surveillance system. Twenty-four open-ended questions were also included to allow participants to provide additional comments or suggestions. The full survey is provided in the supplementary material (see Supplementary File 2).

### Analyses

We described all influenza and COVID-19 cases in the CH-SUR database during the study period. We expressed continuous variables as median and interquartile range, and categorical variables were expressed as counts and percentages. Pearson’s Chi-squared test or the Wilcoxon rank sum tests were used, using a significance threshold set at 0.05.

We assessed the delay to case entry in CH-SUR, defined as the time from hospital admission (or as the time from positive influenza/SARS-CoV-2 test for nosocomial infections) to date of data entry in CH-SUR. We also assessed the completeness of each of the extracted variables, calculated as the proportion of non-missing data for each of the extracted variables among all cases in the database. For COVID-19 cases, we assessed the completeness of data across the nine periods of the pandemic, described across different COVID-19 waves; for influenza we considered five seasons.

For the analysis of the qualitative survey, answers to the perceived importance, relevance, understanding and adaptability of CH-SUR system were expressed as counts and percentages, and by group of stakeholders (PI, research nurses, data scientists/researchers, coordinator/project managers). The responses to the open-ended questions were analysed using natural language processing algorithms to identify keywords ^12,13^. These words were then visualised using a word cloud, with the size of each word reflecting its frequency in the responses. This approach allowed for a visual representation of the prominent themes and concerns expressed by the participants. In addition, a classic qualitative content analysis (QCA)^14^ was conducted on the open-ended responses. The responses were carefully reviewed and categorised into relevant themes and sub-themes. These themes were then analysed and interpreted to identify common patterns, recurring topics, and any noteworthy insights provided by the participants. All analyses were performed using R version 4.2.1 ^15^ using the tidyverse suite (v1.3.1) for data manipulation, and the wordcloud package (v2.6) for text-mining visualizations. The complete analysis pipeline—including data import, cleaning, quantitative evaluation, and qualitative survey processing—is available under an MIT license on GitHub (https://github.com/jsobel1/CH-SUR_quality_analysis) and archived at Zenodo (10.5281/zenodo.15325761).

### Ethical considerations

The CH-SUR system was approved by the Ethics Committee of the Canton of Geneva, Switzerland (CCER 2018– 00577, and CCER 2020–00827). Data collection was also approved by all local ethics committees. For the qualitative survey, all participants were informed about the purpose of the survey, the voluntary nature of their participation, and the confidentiality of their responses.

### Data availability

The datasets presented in this article are not publicly available. The data that support the findings of this study are available from the Federal Office of Public Health, upon reasonable request.

## Results

### Quantitative evaluation of CH-SUR

Between November 2018 and October 2023, 7,375 cases of influenza were reported to the nationwide CH-SUR surveillance system; between March 2020 and October 2023, 49,235 cases of COVID-19 were reported. Characteristics of influenza and COVID-19 patients are described in Table 1. The table shows that COVID-19 patients had a higher proportion of in-hospital mortality (10%) compared to influenza patients (4%), however the COVID-19 case fatality rate decreased over time. Figure 1A shows the cases of influenza reported to CH-SUR for the past five seasons; Figure 1B illustrates the number of COVID-19 cases reported to CH-SUR throughout the duration of the pandemic. For influenza, the 2020/2021 was notable for the near absence of cases reported to CH-SUR, reflecting the impact of infection control and public health measures for COVID-19 on influenza virus circulation.

**Figure 1.**
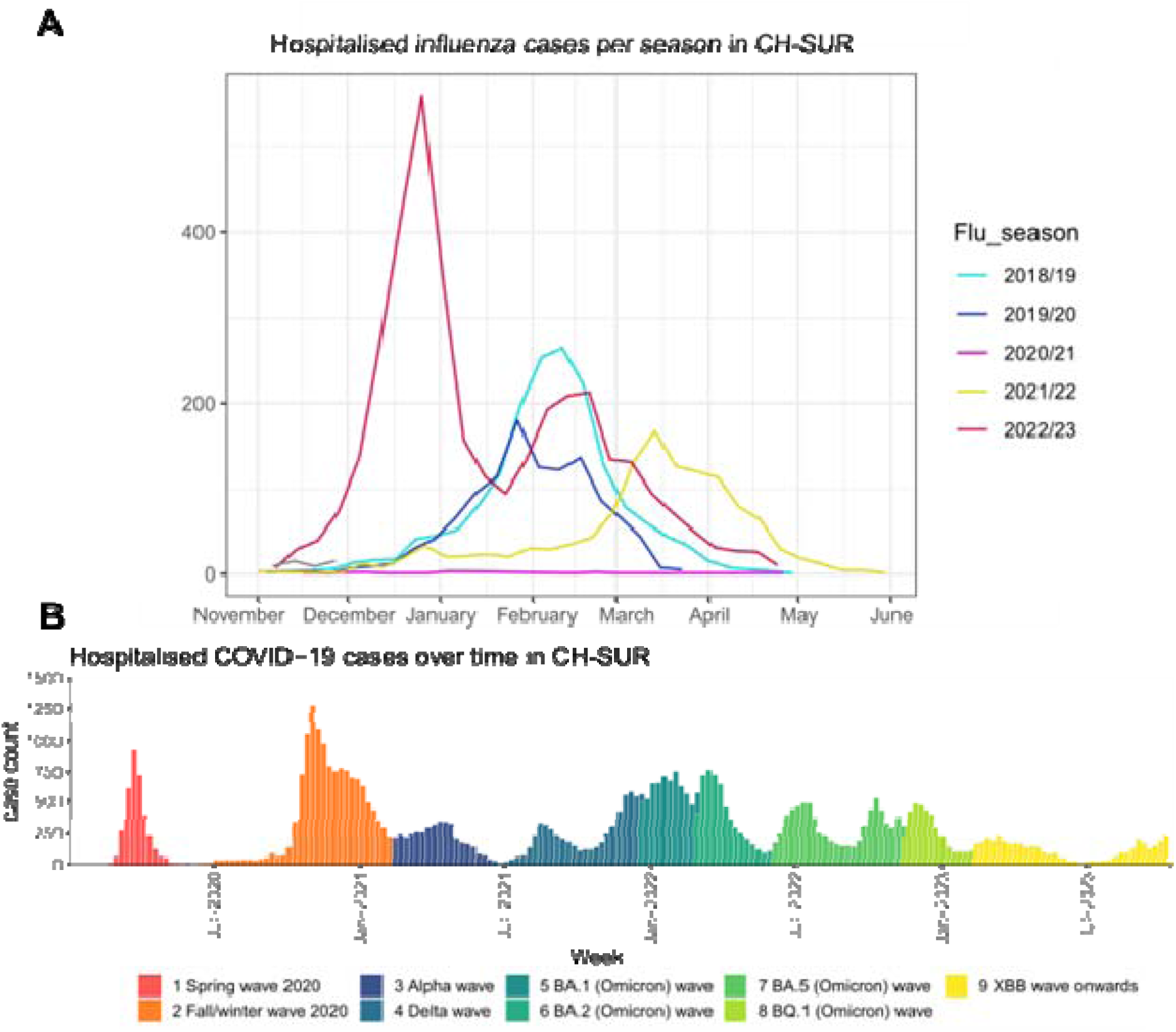
Cases of influenza (A) and COVID-19 (B) reported through the nationwide hospital-based surveillance system in Switzerland between November 2018 and October 2023. The colours in (A) correspond to the respective influenza season; the colours in (B) correspond to the nine different time periods across the COVID-19 pandemic.

**Table 1.**
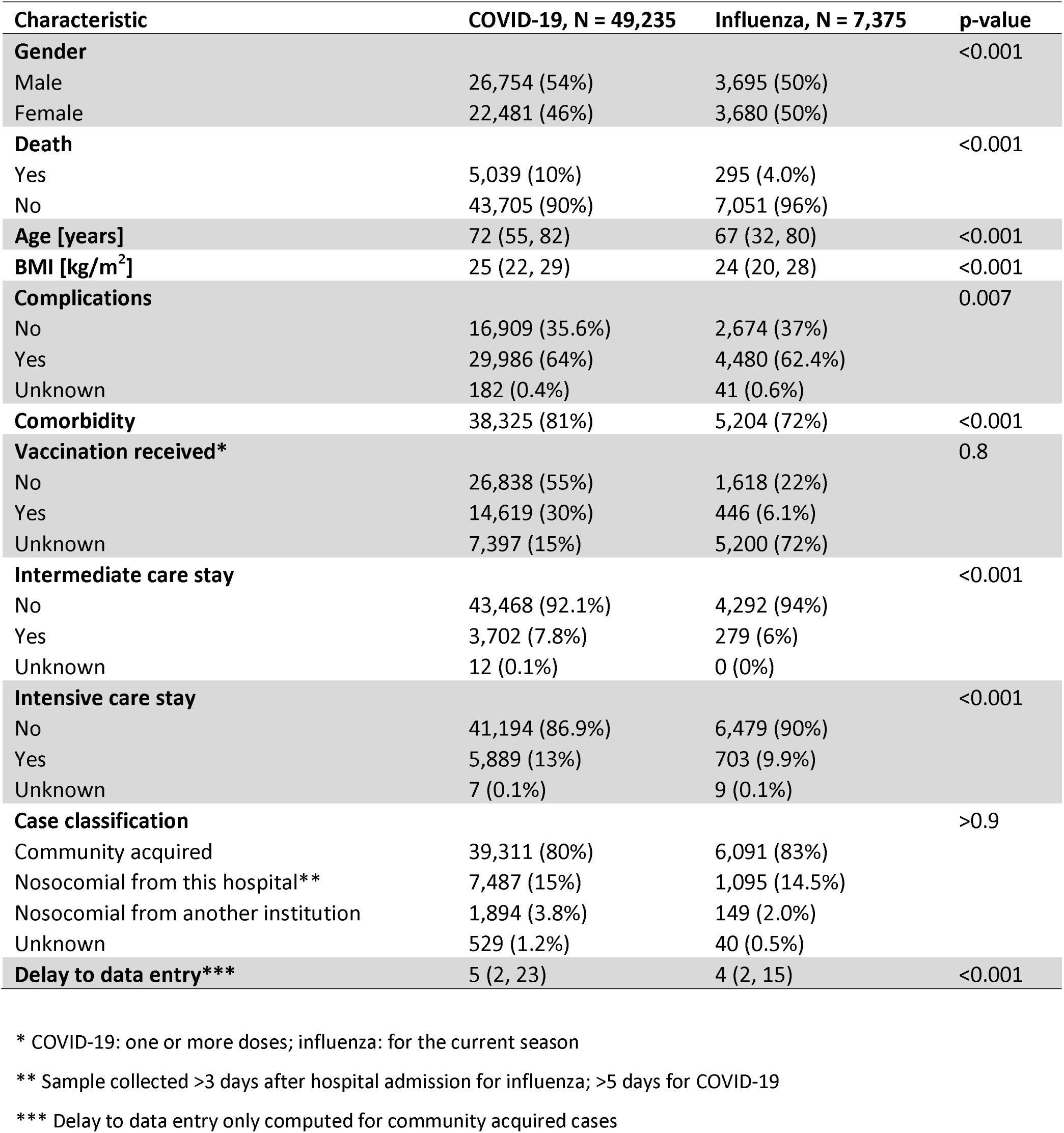
Characteristics of influenza (n=7’375) and COVID-19 (n=49’235) patients reported through the nationwide hospital-based surveillance system in Switzerland between 1 November 2018 and 17 October 2023. Counts and percentages are shown for categorical variables, while medians and interquartile range are shown for continuous variables.

### Delay to case entry

Figure 2 shows the time from hospital admission (or positive influenza /SARS-CoV-2 test for nosocomial infections) to CH-SUR data entry for influenza patients and COVID-19 patients. The median time to case entry for influenza throughout the monitoring period was low (3-8 days from admission, Figure 2A), although the absolute number of influenza cases was also lower as compared to COVID-19. For COVID-19, the median time from admission to case entry in CH-SUR was 5 (IQR:2-23) days and was highest during the initial COVID-19 waves in 2020.

**Figure 2.**
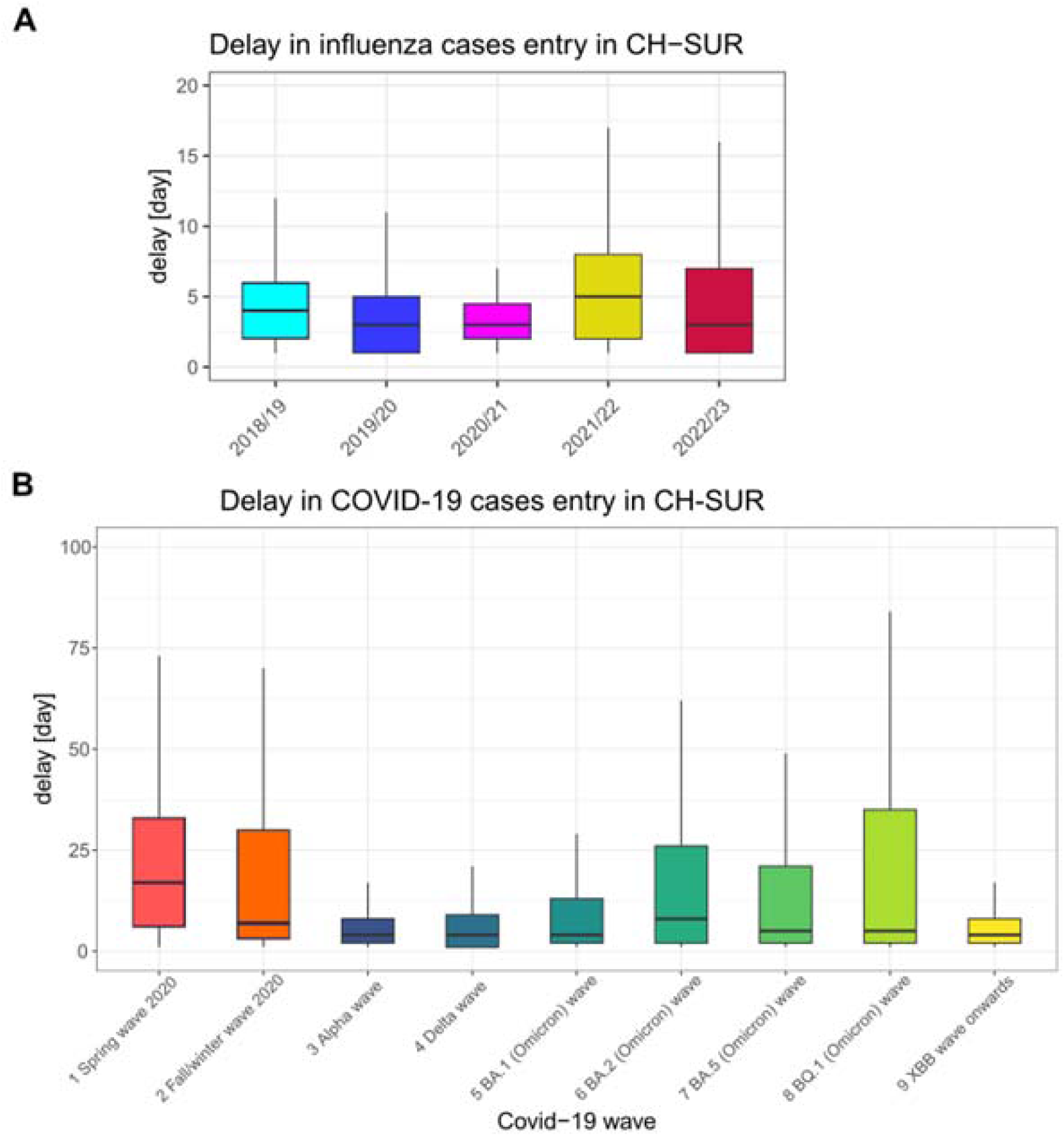
Time from hospital admission (or a positive influenza test/SARS-CoV-2 test for hospital acquired infection) to CH-SUR data entry for influenza patients (A) and COVID-19 patients (B). Each box shows the median and IQR in days. For influenza, the data are shown by influenza season; for COVID-19, the data are shown by nine different time periods across the COVID-19 pandemic. As a reminder: Spring wave 2020 (24 February 2020 to 7 June 2020), 2) Fall/winter wave 2020 (8 June 2020 to 14 February 2021), 3) Alpha wave (15 February 2021 to 20 June 2021), 4) Delta wave (21 June 2021 to 19 December 2021), 5) BA.1 (Omicron) wave (20 December 2021 to 28 February 2022), 6) BA.2 (Omicron) wave (1 March 2022 to 5 June 2022), 7) BA.5 (Omicron) wave (6 June 2022 to 14 November 2022), 8) BQ.1 (Omicron) wave (15 November 2022 to 11 February 2023), and 9) XBB wave onwards (12 February 2023 to 17 December 2023).

### Data completeness

While missing data for vaccination status was low (ranging from 0 to 2.2% of NAs), there was a high percentage of “Unknown” vaccination status, up to 44% in the last period considered. Higher completeness was observed for variables such as comorbidities, intermediate care or intensive care stay, ranging from 0% to 8.6% of NAs (Figures 3A and 3B). The median percentage of complete data was 99.4% across all time intervals and considered variables. With respect to completeness of the components of the CRF, clinical data, including outcome information had the highest proportion of incompleteness (15%) during the last period (Omicron XBB wave onwards), although this likely reflects the fact that patients may have remained hospitalised at the time of the data extraction. Taken together, the time required for data entry and the completeness of the data both improved progressively over time (Figure 2 and 3).

**Figure 3.**
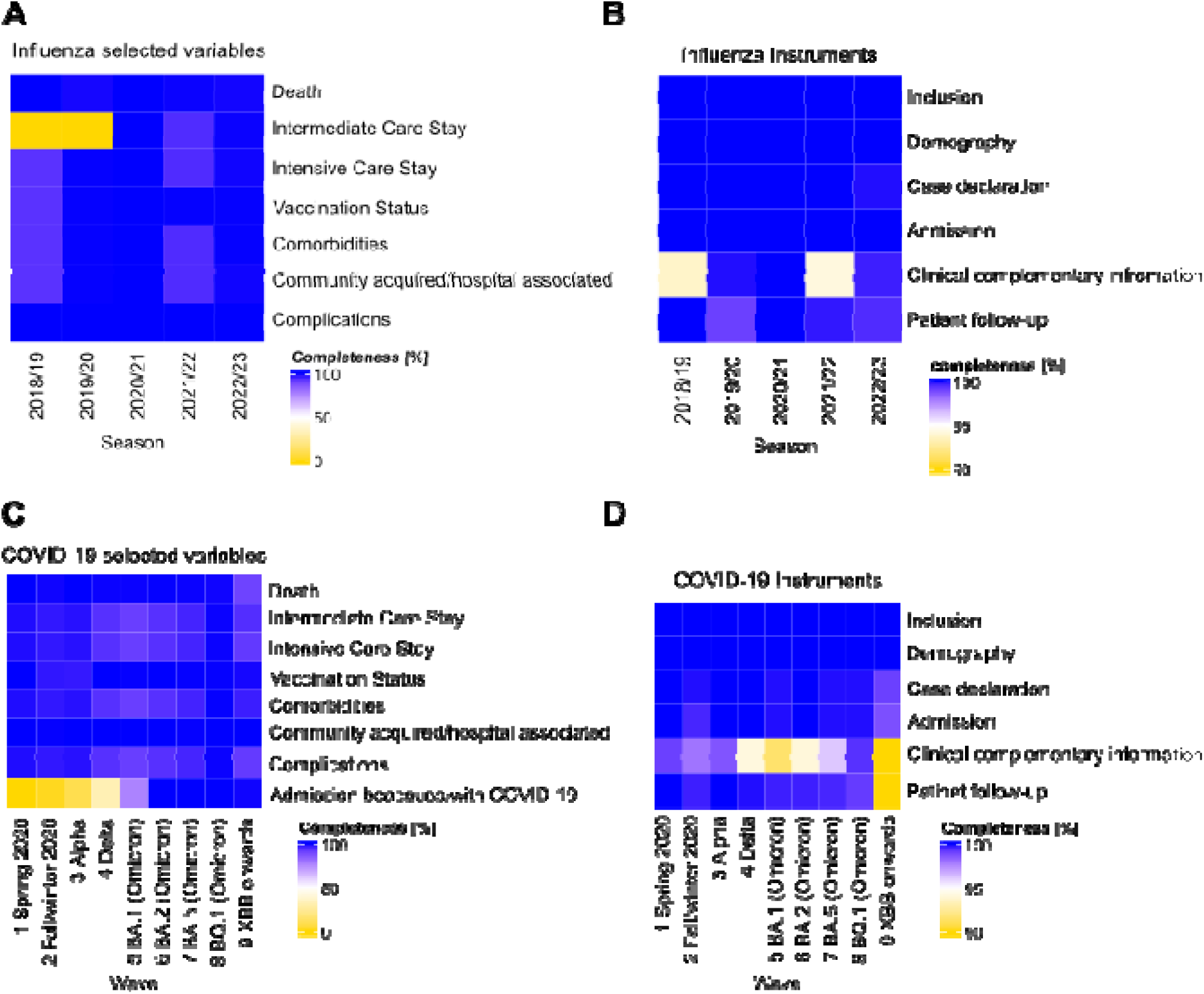
Completeness of data in CH-SUR for influenza (A-B) and COVID-19 patients (C-D). The left panel shows the completeness of the following variables: death, ICU/IMCU stay, comorbidity, complication, reason for admission “because” or “with” COVID-19 and the classification between nosocomial or community acquired, and vaccination; the right panel shows the completeness of the *REDCap* CRF components.

### Qualitative evaluation of CH-SUR

The qualitative survey on CH-SUR was conducted between 1 February 2023 and 1 April 2023. Forty-seven participants responded (Supplementary Figure S3): 13 PIs, 24 study nurses, 6 data scientists/researchers, 4 coordinators/project managers. Most PIs (9/13; 69%) and researchers (5/6; 83%) had been working within the CH-SUR system for more than 2 years. About 40% of nurses had involved in the CH-SUR system for 1 to 2 years (10/24; 41.6%) or more than 2 years (9/24; 37.5%), respectively.

### Stakeholder perceptions

Most CH-SUR stakeholders who participated in the survey judged CH-SUR to be important and relevant; most participants expressed a good or excellent understanding of the system and found it reliable and adaptable to the needs of each respective centre (Figure 4). CH-SUR PIs largely expressed support for expanding CH-SUR to respiratory syncytial virus (RSV) (76.9%) and other diseases (53%). However, they were largely unsupportive (85%) of an expansion of the CH-SUR CRF to include other variables.

**Figure 4.**
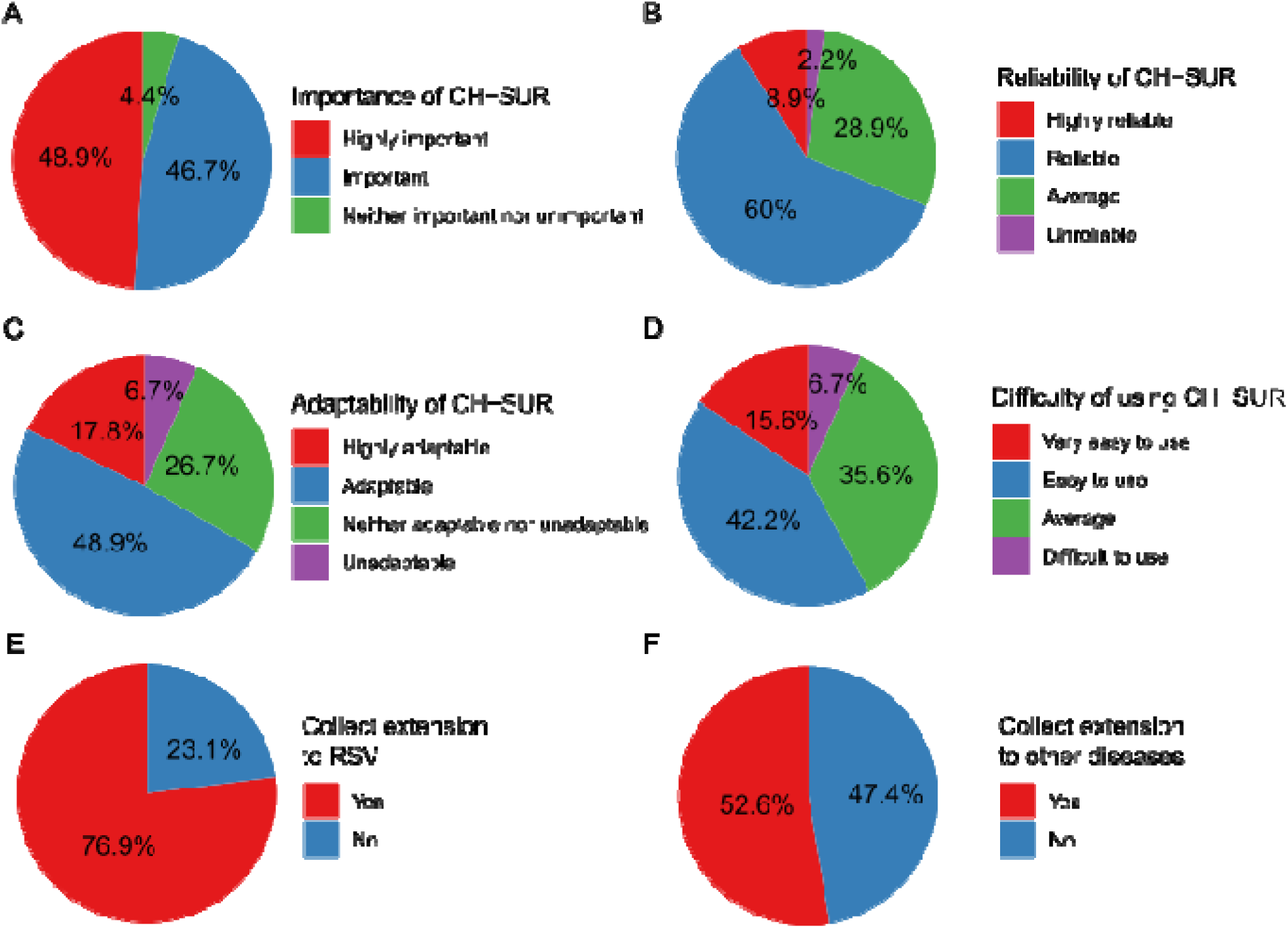
Survey Assessment of CH-SUR Features and Extensions completed by CH-SUR stakeholders. This figure depicts participant responses regarding various aspects of CH-SUR: (A) Perceived importance of CH-SUR (N=45). (B) Perceived reliability of CH-SUR(N=45). (C) Adaptability of CH-SUR in different contexts (N=45). (D) Ease of use of CH-SUR (N=45). (E) Interest in adding Respiratory Syncytial Virus (RSV) monitoring to CH-SUR (N=13). (F) Interest in including monitoring for other diseases in CH-SUR (N=19). Each pie chart represents the distribution of participant opinions on the specified feature or potential extension of CH-SUR.

The survey responses on nurse practice and response timeliness of CH-SUR (Supplementary Figure 4) revealed both strengths in training and follow-up practices, and areas for improvement in response timeliness and efficiency.

The open-ended questions of the survey identified several challenges faced by CH-SUR stakeholders. The first related to personnel and the ability to hire nurses to enter data into the CH-SUR system, as well as collaborators having to work overtime, particularly during the early stages of the COVID-19 pandemic. This was closely linked to financial challenges in resource allocation and budgeting for CH-SUR activities. CH-SUR stakeholders reported challenges related to the electronic CRF, including pop-up messages, violation rules, and potential issues with missing or incorrect values. The automation of data entry within the CRF also presented a substantial challenge as only some of the larger institutions had the capacity to partially automate the extraction of some of the variables from patients’ electronic health records. The text mining and word cloud analysis of responses to the open-ended questions in the survey showed that CH-SUR is centred on "data" and “cases”. Among frequently cited themes, we observed the terms "nurses", "weekly", "report" as well "automation", "COVID-19", "surveillance", "data quality", "manpower", and "workload" (Supplementary Figure 5).

## Discussion

We conducted an evaluation of the nationwide hospital-based surveillance for influenza and COVID-19 in Switzerland from November 2018 to October 2023, following CDC’s guidelines for evaluating public health surveillance systems^10^. In addition, we conducted a qualitative stakeholder survey in February-April 2023. Initially established for surveillance of influenza, the expanded surveillance system has enabled Swiss health authorities to monitor epidemiological trends in hospitalised influenza and COVID-19 patients across Switzerland, to quantify the respective disease burden and to assess healthcare demands over time.

Within one month of the report of the first confirmed case of COVID-19 in Switzerland, the surveillance system was expanded to include what was then a novel respiratory disease. This demonstrates the importance of an established network with an agile, adaptable surveillance system and motivated participating hospitals to include other diseases beyond the initial intention. Further expansion of CH-SUR to include RSV, for example, may be considered in the future. In terms of preparedness for future pandemics, it also demonstrates the time and resource advantages of adapting existing surveillance systems, rather than having to establish a de novo surveillance system ^16^. The flexibility of the system was also demonstrated by its ability to capture COVID-19 vaccination details as COVID-19 vaccines were introduced, and COVID-19 treatments as different therapeutics were either repurposed to treat COVID-19 or developed specifically for COVID-19 ^17^. The three-fold increase in the number of participating centres after the onset of the pandemic is a testament to the system’s ability to be implemented across healthcare settings: tertiary and non-tertiary care settings; adult and paediatric settings. Completeness and timeliness are two additional indicators of the quality of the surveillance system. Our quantitative analyses demonstrated a high level of completeness for variables within the CRF and timely reporting of cases throughout most of the pandemic.

CH-SUR strengthened Switzerland’s surveillance landscape by capturing data on hospitalised cases, complementing systems focused on outpatient care and community circulation: while Sentinella monitors influenza-like illness and circulating respiratory viruses in primary care and wastewater surveillance tracks viral circulation at the population level, CH-SUR provided critical data on the severity and healthcare burden of COVID-19 and influenza, supporting public health decision-making. Beyond the primary public health objectives of the surveillance system, the collection of data on hospitalised patients also allowed important scientific questions to be addressed. This included identifying risk factors for severe COVID-19 outcomes ^18^; an evaluation of COVID-19 mortality over time ^19^; a comparison of clinical outcomes among community-acquired influenza and COVID-19 and a comparison of community acquired and healthcare-associated COVID-19 ^20,21^.

Our qualitative evaluation of the CH-SUR surveillance system provided further insights from CH-SUR stakeholders. It highlighted that the expansion of the surveillance system to a novel disease was not without notable challenges ^22^. One difficulty stemmed from the absolute number of COVID-19 cases requiring hospital-level care at each of the participating centres especially during the first pandemic phase was extremely high, and the increased scale and speed with which data needed to be reported. Although not directly shown in our analysis, it has been shown that the protracted nature of the COVID-19 pandemic has led to burnout among nurses and other healthcare workers due to overtime work, lack of holidays, and understaffing ^23,24,25^. This underscores the need for strategic human resource planning and sufficient financial resources to ensure the surveillance system can achieve its stated objectives. Future cost-effectiveness analyses of CH-SUR may also help to ‘right-size’ the surveillance system.

Data entry for CH-SUR currently relies on study nurses retrieving data from patients’ (mostly computerised) medical files and manually entering these data into the CRF. Unlike centralised systems in other countries, such as France, Australia and New Zealand ^26^, each centre participating in CH-SUR has its own system for managing patients’ medical data. In alignment with the digitalisation strategy and automatization of surveillance led by Swissnoso and FOPH, automating data extraction and management may address some of the challenges identified through our evaluation. This would not resolve the need for careful interpretation and clinical judgement of some data, and there would be a challenge in ensuring interoperability between different systems and databases. However, pilot studies on automated surveillance are ongoing in Switzerland, and CH-SUR may be able to benefit from the technological advances used in these initiatives. The FAIR (Findable, Accessible, Interoperable, Reusable) principles’ application to biomedical databases is instrumental in enhancing the value and utility of data ^27^ and it would be important to ensure that any automated CH-SUR system adheres to these principles.

This study has some limitations. First, the true number of hospitalised influenza and COVID-19 cases not reported to CH-SUR remains unknown, potentially affecting the accuracy of the findings. Further, the qualitative assessment was based on a convenience sample, which may not be representative of all CH-SUR stakeholders, thus limiting the generalizability of the results. These factors collectively highlight the need for caution in interpreting the outcomes of this analysis.

## Conclusions and perspectives

Overall, the nationwide hospital-based surveillance for influenza was successfully adapted to COVID-19 ^28^ and has provided important, actionable information on epidemiological trends in hospitalised patients across Switzerland. This system has been instrumental in understanding the disease burden among hospitalised patients and healthcare demand, serving as a crucial resource for guiding evidence-based public health decisions and preparing the Swiss health system for future respiratory virus outbreaks. Our evaluation highlighted the significance and relevance of this system among CH-SUR stakeholders, with identified challenges serving as opportunities for improvement towards a sustainable and efficient respiratory disease surveillance system in Switzerland.

Starting in January 2024, significant changes were implemented in the CH-SUR system. Throughout the pandemic, the system had been financed through temporary credits by FOPH, which were significantly reduced in 2024. The network was thus first reduced to six hospitals from January 2024 and discontinued on 1 September 2024, due to financial constraints. Looking ahead, further developments in collection capabilities through increased automation for a future national hospital surveillance system are recommended. This includes the implementation of the CH-SUR statistics on the Infectious Disease Dashboard of the FOPH and the extension of surveillance to other diseases, such as RSV. These advancements aim to enhance surveillance sustainability and resilience, and data accuracy and accessibility, providing comprehensive insights into multiple infectious diseases. The implementation of automated processes and user-friendly dashboards will facilitate real-time monitoring and analysis, ultimately strengthening public health surveillance and response strategies. By embracing these innovations, Switzerland can ensure a robust and adaptable surveillance system capable of addressing current and future public health challenges ^29^.

## Data Availability

CH-SUR data can be accessed upon request through the CH-SUR scientific committee's approval process.

## Acknowledgements

We would like to thank the members of the CH-SUR collaborative network who helped conduct the study in each of the study sites. In particular, Mohamed Abbas, Jason Toko, Daniel Texiera, Michèle Steiner, Marianne Rousseau-Schadegg, Anne-Flore Combaz, Audrey Aymon, Carlo Balmelli, Manuel Battegay, Christoph Berger, Sara Bernhard-Stirnemann, Elfriede Berwarth, Julia Bielicki, Michael Büttcher, Alexia Cusini, Lauro Damonti, Philipp Kaiser, Stefan Keller, Henrik Köhler, Elia Lo Priore, Yvonne Nussbaumer, Matthaios Papadimitriou, Felix Reichlin, Thomas Riedel, Susanne Rüfenacht, Hanna Schmid, Markus Schneemann, Laurence Senn, Reto Stocker, Nicolas Troillet, Sarah Tschudin-Sutter, Cathy Voide, Danielle Vuichard Gysin, Konstanze Zoehrer, Franziska Zucol. We would also like to thank Céline Gardiol and Jasmin Vonlanthen, Ornella Luminati, Carolina Agop Nersesian, Carla Grolimund, Fabienne Krauer, Mirjam Mäusezahl, and Katrin Schneider from the Federal Office of Public Health of Switzerland and Maroussia Roelens, and Amaury Thiabaud from the University of Geneva. We acknowledge the contributions of the Clinical Research Center, Geneva University Hospitals and Faculty of Medicine, Geneva. We acknowledge that this study was made possible through the funding and supervision provided by FOPH and the Swiss National Science Foundation (grant no 31CA30_196270).

## Supplementary material

Supplementary file 1: CH-SUR case report form and codebook, version 2023.

Supplementary file 2: CH-SUR evaluation survey

**Supplementary Figure 1.**
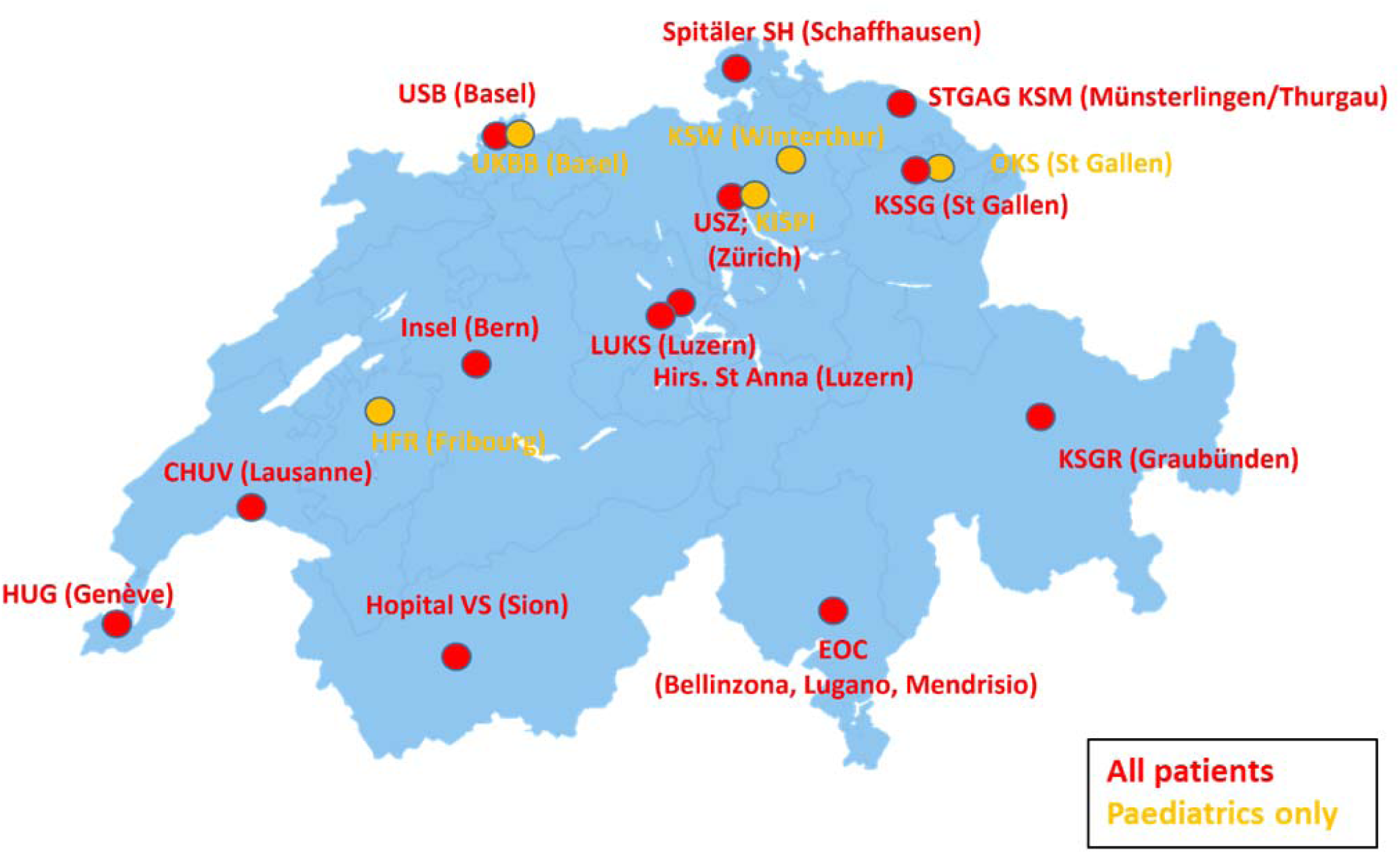
Map of CH-SUR Hospital partners over Switzerland., KSSG - Kantonsspital St. Gallen, HUG - Hôpitaux Universitaires de Genève, UKBB - Universitäts-Kinderspital Basel (University Children’s Hospital Basel), LUKS - Luzerner Kantonsspital, KISPI USZ - Kinderspital Zürich (University Children’s Hospital Zurich), PED KSW - Kinderspital Winterthur (Children’s Hospital Winterthur), EOC - Ente Ospedaliero Cantonale (Cantonal Hospital Organization), PED HFR - Pédiatrie Hôpital fribourgeois (Pediatrics, Fribourg Cantonal Hospital), PED OKS - Pédiatrie Ospedale cantonale di San Gallo (Pediatrics, Cantonal Hospital of St. Gallen), HVS - Hôpital du Valais, STGAG - Spital Thurgau AG, USZ - Universitätsspital Zürich (University Hospital Zurich), CHUV - Centre Hospitalier Universitaire Vaudois, USB - Universitätsspital Basel (University Hospital Basel), SSH - Spitäler Schaffhausen, Hirslanden LU - Hirslanden Klinik St. Anna Luzern, INSEL - Inselspital (University Hospital of Bern)

**Supplementary Figure 2.**
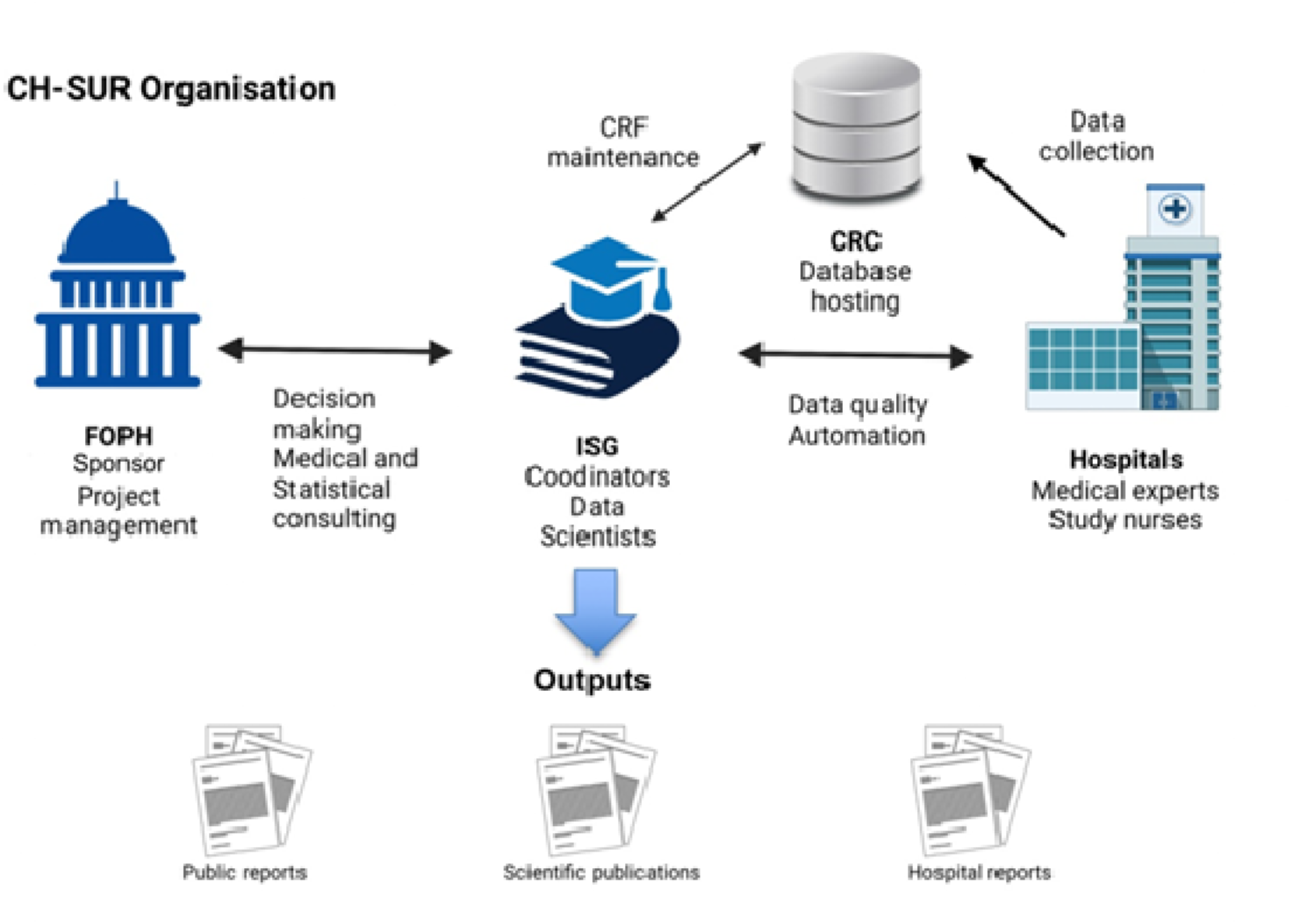
Organisation of nationwide hospital-based surveillance system for influenza and COVID-19 in Switzerland (CH-SUR). FOPH: Federal Office of Public Health; ISG: Institut de Santé Globale, Université de Genève; CRC: Centre de recherche clinique, Geneva University Hospitals; CRF: Case report form * As of 2024, several tasks were internalised at the FOPH, including reporting and data science functions.

**Supplementary Figure 3.**
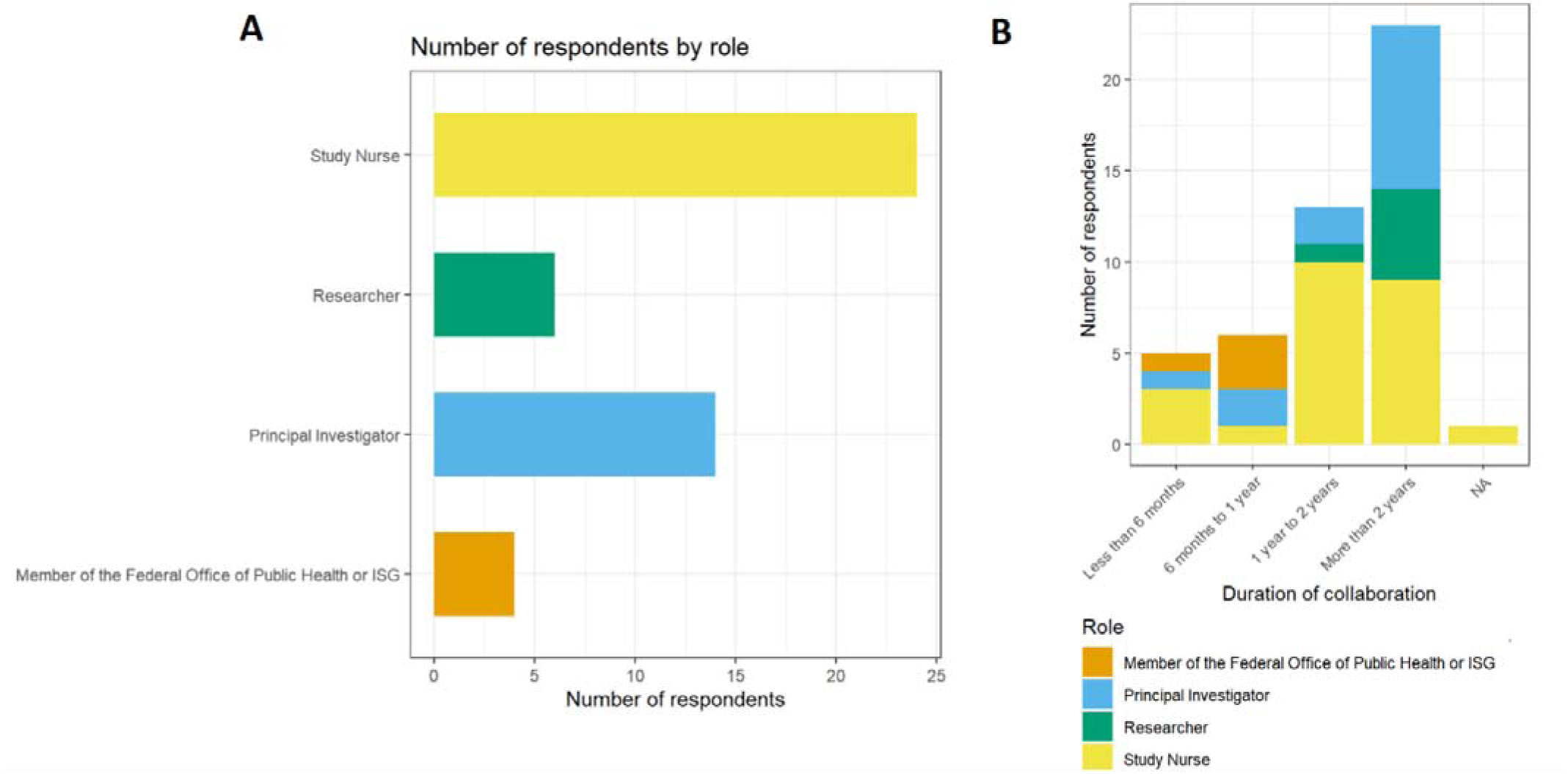
Roles within CH-SUR (A) and duration of collaboration (B) among qualitative survey participants CH-SUR.

**Supplementary Figure 4.**
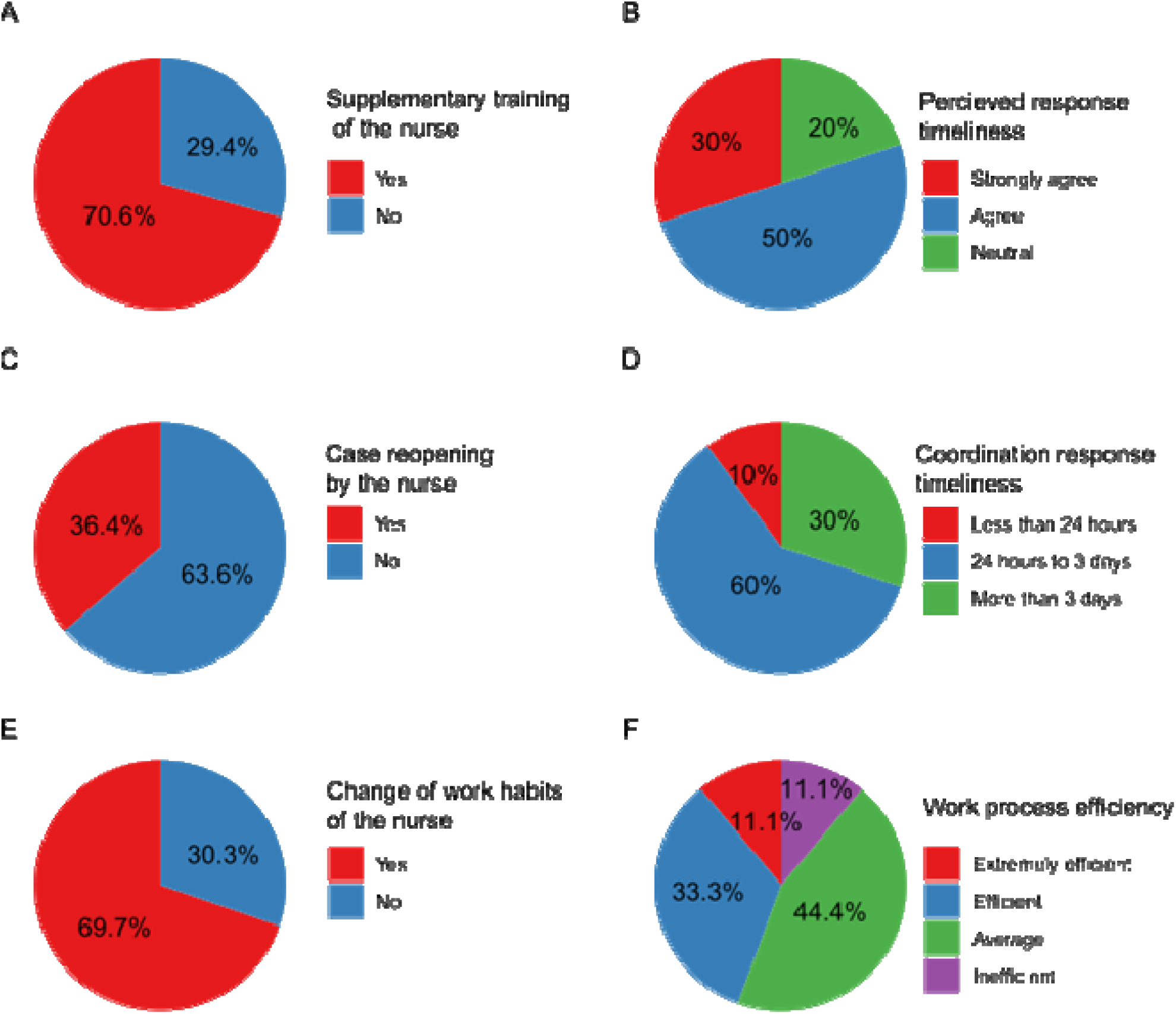
Survey responses on nurse’s practices and response timeliness. (A) Additional training of nurses (N=34). (B) General satisfaction with response timeliness from CH-SUR (N=10). (C) Case follow-up practices by nurses (N=33). (D) Specific timeframes for CH-SUR responses (N=10). (E): Change of work habits by nurses (N=33). (F) Assessment of the efficiency of CH-SUR’s work processes (N=9).

**Supplementary Figure 5.**
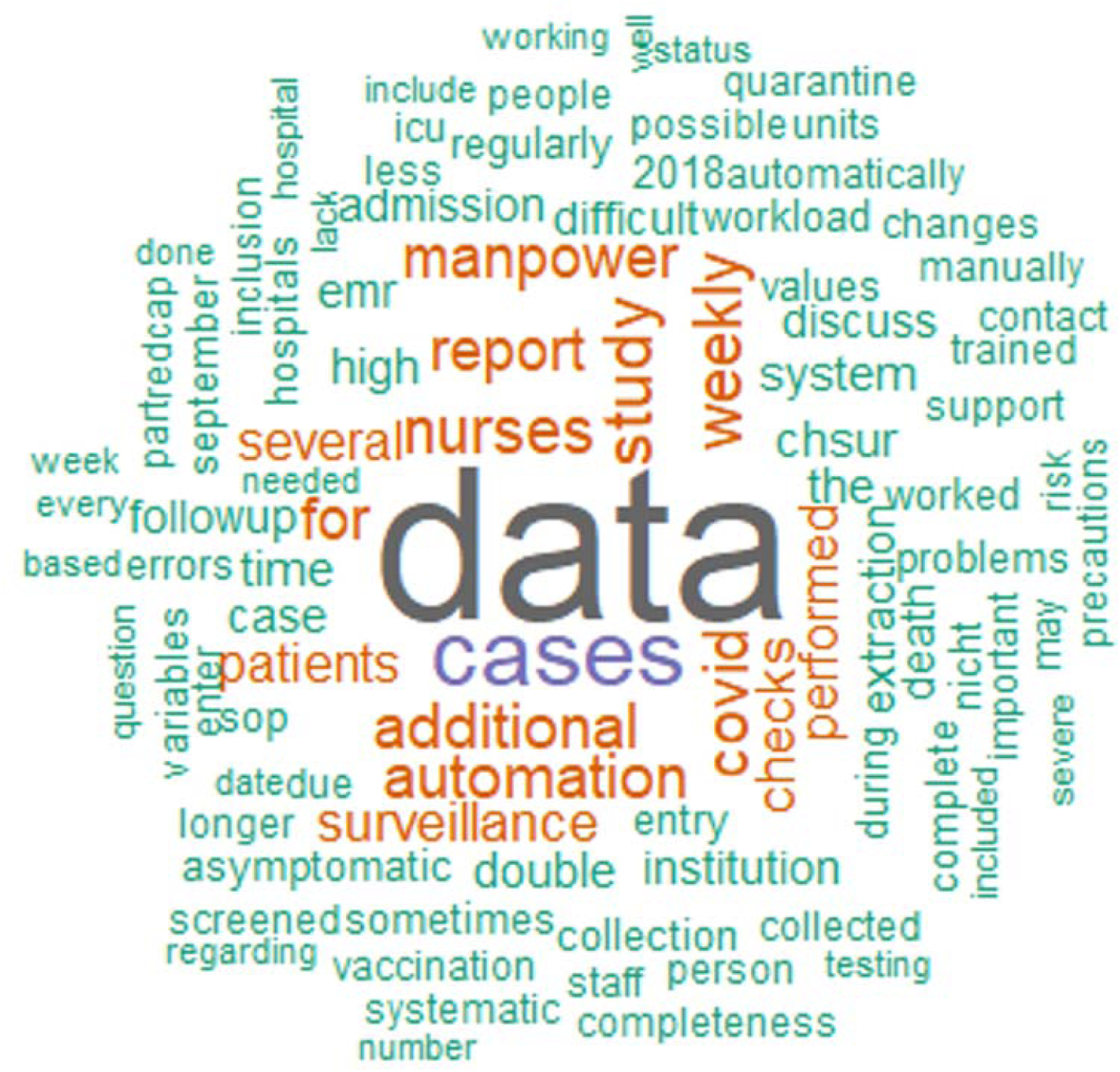
Results of text mining and word cloud analysis of open-ended questions in CH-SUR survey. The size of the words is proportional to the occurrence of the word in the respondents’ answers.

## References

1. Abbas M, Zhu N, Mookerjee S, et al. Hospital-onset COVID-19 infection surveillance systems: a systematic review. Journal of Hospital Infection. 2021;115:44–50.

2. Marcenac P, McCarron M, Davis W, et al. Leveraging international influenza surveillance systems and programs during the COVID-19 pandemic. Emerging infectious diseases. 2022;28(Suppl 1):S26.

3. Fougerolles de, Damm O, Ansaldi F, et al. National influenza surveillance systems in five European countries: a qualitative comparative framework based on WHO guidance. BMC Public Health. 2022;22(1):1–13.

4. Quiros-Gonzalez V, Rodriguez-Perez P, Haro-Perez AM, Jimenez-Rodriguez MM, Maderuelo-Fernandez J Angel, Eiros JM. Real-time surveillance systems: applicability for the control of influenza in acute care. Influenza and other respiratory viruses. 2020;14(3):331–339.

5. Harris PA and T Robert, Thielke R, Payne J, Gonzalez N, Conde JG. Research electronic data capture (REDCap)—a metadata-driven methodology and workflow process for providing translational research informatics support. Journal of biomedical informatics. 2009;42(2):377–381.

6. New Coronavirus 2019-nCoV: first confirmed case in Switzerland. Accessed June 5, 2024. https://www.bag.admin.ch/bag/en/home/das-bag/aktuell/medienmitteilungen.msg-id-78233.html

7. Sousa M, Roelens M, Fricker B, et al. Risk factors for severe outcomes for COVID-19 patients hospitalised in Switzerland during the first pandemic wave, February to August 2020: prospective observational cohort study. Swiss medical weekly. 2021;151:w20547.

8. Thiabaud A, Iten A, Balmelli C, et al. Cohort profile: SARS-CoV-2/COVID-19 hospitalised patients in Switzerland. Swiss medical weekly. 2021;151:w20475.

9. Swiss Federal office of public health. Monthly COVID-19 situation in Switzerland. Published online June 2023.

10. Centers for Disease Control and Prevention (CDC), Updated Guidelines for Evaluating Public Health Surveillance Systems, Recommendations from the Guidelines Working Group, 2001

11. Shu Y, McCauley J. GISAID: Global initiative on sharing all influenza data – from vision to reality. Eurosurveillance. 2017;22(13). doi:10.2807/1560-7917.es.2017.22.13.30494

12. Fellows I. Wordcloud: Word Clouds.; 2018. https://CRAN.R-project.org/package=wordcloud

13. Feinerer I, Hornik K, Meyer D. Text Mining Infrastructure in R. Journal of Statistical Software. 2008;25(5):1–54. doi:10.18637/jss.v025.i05

14. Schreier M. Qualitative Content Analysis. In: The SAGE Handbook of Qualitative Data Analysis. SAGE Publications, Inc.; 2014:170–183. Accessed June 5, 2024. 10.4135/9781446282243.n12

15. Team RC. R: A Language and Environment for Statistical Computing. R Foundation for Statistical Computing; 2023. https://www.R-project.org/

16. Ricks PM, Njie GJ, Dawood FS, et al. Lessons learned from CDC’s global COVID-19 Early Warning and Response Surveillance system. Emerging Infectious Diseases. 2022;28(Suppl 1):S8.

17. Estill J, Venkova-Marchevska P, Günthard HF, et al. Treatment effect of remdesivir on the mortality of hospitalised COVID-19 patients in Switzerland across different patient groups: a tree-based model analysis. Swiss Medical Weekly. 2023;153(8):40095. doi:10.57187/smw.2023.40095

18. Frohlich GM, Kraker D, Abbas M, et al. Hospital outcomes of community-acquired COVID-19 versus influenza: Insights from the Swiss hospital-based surveillance of influenza and COVID-19. Eurosurveillance. 2022;27(1):2001848.

19. Roelens M, Martin A, Friker B, et al. Evolution of COVID-19 mortality over time: results from the Swiss hospital surveillance system (CH-SUR). medRxiv. Published online 2021:2021–09.

20. Portmann L, Kraker de, Frohlich G, et al. Hospital Outcomes of Community-Acquired SARS-CoV-2 Omicron Variant Infection Compared With Influenza Infection in Switzerland. JAMA Network Open. 2023;6(2):e2255599–e2255599.

21. Grant RL, Sauser J, Atkinson A, et al. Comparison of clinical outcomes over time of inpatients with healthcare-associated or community-acquired coronavirus disease 2019 (COVID-19): A multicenter, prospective cohort study. Infection Control & Hospital Epidemiology. 2023;45(1):75–81. doi:10.1017/ice.2023.143

22. Bloom DE, Cadarette D. Strengthening the Global Response to Infectious Disease Threats in the Twenty-First Century, with a COVID-19 Epilogue. In: Global Health. Cambridge University Press; 2021:51-75. Accessed June 13, 2024. 10.1017/9781108692137.004

23. Sullivan D, Sullivan V, Weatherspoon D, Frazer C. Comparison of nurse burnout, before and during the COVID-19 pandemic. Nursing Clinics. 2022;57(1):79–99.

24. Lasater KB, Aiken LH, Sloane DM, et al. Chronic hospital nurse understaffing meets COVID-19: an observational study. BMJ Quality \& Safety. 2021;30(8):639–647.

25. Rivas N, Lopez M, Castro MJ, et al. Analysis of burnout syndrome and resilience in nurses throughout the COVID-19 pandemic: a cross-sectional study. International journal of environmental research and public health. 2021;18(19):10470.

26. Lee J, Park YT, Park YR, Lee JH. Review of National-Level Personal Health Records in Advanced Countries. Healthcare Informatics Research. 2021;27(2):102–109. doi:10.4258/hir.2021.27.2.102

27. Queralt-Rosinach N, Kaliyaperumal R, Bernabe CH, et al. Applying the FAIR principles to data in a hospital: challenges and opportunities in a pandemic. Journal of biomedical semantics. 2022;13(1):1–19.

28. World Health Organization, others. WHO Consultation to Adapt Influenza Sentinel Surveillance Systems to Include COVID-19 Virological Surveillance: Virtual Meeting, 6--8 October 2020. World Health Organization; 2022.

29. World Health Organization, others. COVID-19 Strategic Preparedness and Response Plan: Monitoring and Evaluation Framework, 11 May 2021. World Health Organization; 2021.

